# Pilot study of a ketogenic diet in bipolar disorder

**DOI:** 10.1101/2023.05.28.23290595

**Authors:** Nicole Needham, Iain H Campbell, Helen Grossi, Ivana Kamenska, Benjamin P Rigby, Sharon A Simpson, Emma McIntosh, Pankaj Bahuguna, Ben Meadowcroft, Frances Creasy, Maja Mitchell-Grigorjeva, John Norrie, Melissa C Gibbs, Ailsa McLellan, Cheryl Fisher, Tessa Moses, Karl Burgess, Rachel Brown, Michael Thrippleton, Harry Campbell, Daniel J Smith

## Abstract

**Background:** Recent evidence from case reports suggests that a ketogenic diet may be effective for bipolar disorder. To date, no clinical trials have been conducted.

**Aims:** To assess the recruitment and feasibility of a ketogenic diet intervention in bipolar disorder.

**Methods:** Euthymic individuals with bipolar disorder were recruited to a 6-8 week trial of a modified ketogenic diet and a range of clinical, economic and functional outcome measures were assessed.

**Results:** Of 27 recruited participants, 26 commenced and 20 completed the modified ketogenic diet at 6-8 weeks. The completeness of the outcomes dataset was 95% for daily ketone measures, 95% for daily glucose measures and 95% for daily Ecological Momentary Assessment of symptoms during the intervention period. Mean daily blood ketone readings were 1.3 mmol/L (SD= 0.77, Median = 1.1), and 91% of all readings indicated ketosis indicating a high degree of adherence to the diet. Over 91% of daily blood glucose readings were within normal range with 9% indicating mild hypoglycaemia. Eleven minor adverse events were recorded, including fatigue, constipation, drowsiness and hunger. One serious adverse event was reported (euglycemic ketoacidosis in a participant taking *SGLT2*-inhibitor medication).

**Conclusions:** The recruitment and retention of euthymic individuals with bipolar disorder to a 6-8 week ketogenic diet intervention was feasible, with high outcome measure completion rates. The majority of participants reached and maintained ketosis and adverse events were generally mild and modifiable. A future randomised controlled trial is now warranted.

**Study Registration Number:** ISRCTN61613198

## Introduction

Bipolar disorder is a serious mental illness with a lifetime risk of 1-2% (1). The global burden of bipolar disorder, measured in disability-adjusted life years (DALYs), rose by 14.9% over the period 2005-15 (2). It usually begins in early adulthood and has considerable economic and societal impact. However, the underlying pathophysiology remains poorly understood and current treatment and prevention strategies are suboptimal, with a need to identify effective new interventions.

Abnormal glucose metabolism is common in people with bipolar disorder, with an increased prevalence of insulin resistance and type 2 diabetes (3,4). It is possible that hyperinsulinemia may adversely affect the production of energy by mitochondria in the brain (5-7). Ketones can provide an alternative energy source to glucose, which may bypass this process and allow for more stable levels of energy production (8).

The ketogenic diet is a safe and effective therapy for seizure reduction in children with drug-resistant epilepsy, evidenced by a Cochrane review of randomised controlled trials and almost a century of clinical use (9,10). The diet’s most significant effect is that it induces ketosis wherein mitochondria in the body and brain preferentially oxidise fatty acids to produce ketones (beta hydroxybutyrate and acetoacetate) and then use these as a source of energy (instead of glucose). This metabolic shift reduces seizures by 50% in around half of children with drug-resistant epilepsy (11). There are clinical parallels between bipolar disorder and epilepsy, which could indicate similarities in underlying disease processes (12).

Case series have suggested benefits of a ketogenic diet in several psychiatric disorders, including bipolar disorder (13) and schizophrenia (14). Analyses of data from online bipolar disorder forums (165 people with bipolar disorder adhering to a ketogenic diet) found that 56% reported either remission of symptoms or significant mood stabilisation (4). Recent narrative reviews have identified that a ketogenic diet may impact on metabolic and biochemical features of bipolar disorder, including: reduction of oxidative stress; improved mitochondrial function and biogenesis; improved glutamate/GABA transmission; and reductions of intracellular sodium and calcium (15).

In this pilot study we aimed to assess recruitment, acceptability and feasibility of the intervention, and completion of relevant outcome measures. Secondary objectives (to be reported in a separate publication) were to assess the relationships between biochemical, metabolomic and brain imaging biomarkers with clinical and functional outcomes.

## Methods

### Design, approvals and consent

This was a single-group non-randomised interventional pilot study with no control arm. The authors assert that all procedures contributing to this work comply with the ethical standards of the relevant national and institutional committees on human experimentation and with the Helsinki Declaration of 1975, as revised in 2008. The study received a favourable ethical opinion from the South East Scotland Research Ethics Committee 02, and was approved by NHS Lothian Research and Development. Sponsorship was provided by the Academic and Clinical Central Office for Research and Development (ACCORD). Written informed consent was obtained from all subjects. The study was prospectively registered at isrctn.com under the registration number ISRCTN61613198 on 2 March 2022.

### Study participants

Participants were recruited through Bipolar Scotland via email circulars, local groups, and targeted social media. Eligible participants were those with a diagnosis of bipolar disorder, as per the Diagnostic and Statistical Manual of Mental Disorders (DSM-4) diagnostic criteria, who had been clinically euthymic for 3 months (defined as no episodes of depression or hypomania/mania). They had to be aged 18-70, with the upper age limit set to improve the comparability of MRI head scans. Participants needed to have a sufficient understanding of English, and to be based in Scotland. Initial exclusion criteria included: pregnancy, breastfeeding or planning to become pregnant within 3 months; active substance misuse; use of a ketogenic diet within 2 months; following a vegan diet; admission to hospital within 3 months; involvement in any other research; inability to complete baseline assessments; liver, kidney, or cardiovascular disease; and severe hyperlipidaemia. The following exclusion criteria were added during the study period: diabetes; training for or undertaking very high energy requirement activities; and significant changes to psychotropic medication, planned or within 3 months. Reasons for these additional exclusion criteria are described below.

### Participant assessments and data collection

Pre-recruitment appointments included an explanation of the study to potential participants and checking of eligibility criteria. At baseline appointments, participants received detailed information about establishing and maintaining a ketogenic diet, including potential risks and instructions on how to complete monitoring. They were asked to complete and return a 3 day food diary and a pre ketogenic diet information sheet (see supplementary data) beforehand to aid planning and personalisation of the diet.

At baseline and at 6-8 week follow up assessments, medical and medication history were reviewed, blood pressure and body mass index (BMI) were measured, and a diagnostic interview was completed. Participants completed a range of mental health measures (Affective Lability Scale 18; Beck’s Depression Inventory; Young Mania Rating Scale). Quality of life measures included a Within Trial Resource Use Questionnaire (WTRUQ; developed to identify key health and social care resource use in addition to personal household expenditure on food and beverages and employment/absenteeism information); the EuroQol 5D quality of life instrument (16); and Work Productivity and Activity Impairment Questionnaire (WPAIQ; tailored). Fasting venepuncture and MRI brain scans were completed before and after the intervention. Questionnaires were either completed on paper, or later entered into a secure online platform by participants. Face-to-face contacts took place at the Clinical Research Facility (CRF) and imaging department at the Royal Infirmary of Edinburgh (RIE).

Participants were asked to measure daily capillary readings of glucose and ketones on a KetoMojo device. Initial participants texted daily readings to the research team, but they and later participants were subsequently able to sync readings via bluetooth to an app on their phone, and provide the research team permission to access the results via an online platform. Participants also completed daily ecological momentary assessments (EMAs) of anxiety, mood, energy, impulsivity and speed of thought. These were initially sent by text, with an online data collection tool trialled for the final 2 participants. Participants also completed continuous accelerometry for a 9 week period, using 3 consecutively worn *AX3* actigraph devices for 3 weeks each.

All participants were offered weekly remote contact with a dietitian during the intervention period, with additional contacts as required. They had the contact details of a psychiatrist on the research team, who was available to address concerns about their mental state or other aspects of the study.

Post-intervention, 15 participants and 4 health professionals who were involved in the study took part in a semi-structured interview as part of a process evaluation (reported separately).

### Intervention

The modified ketogenic diet was used, with estimated energy requirements of approximately 60-75% from fat, 5-7% from carbohydrate and additional calories from protein (11). Unsaturated fats were encouraged to minimise the risk of increased blood cholesterol and triglycerides. Individual macronutrient percentages were altered over time based on a range of factors, including levels of ketosis. This modified ketogenic diet works best if participants ingest as near to their daily energy requirements as possible. Where weight loss was desired and safe, an energy deficit was prescribed to encourage the use of body fat as an additional ketone source. Participants were established on the intervention for a 6-8 week period, including a brief adaptation period. Medical treatment from regular care teams continued as standard. The modified ketogenic diet information sheet specified desirable blood ketones levels as 1–4 mmol/L and glucose levels as 4-7.8 mmol/L (see Supplementary Materials).

Support during the intervention period included confirming participants’ understanding of the intervention, supporting adherence to the diet, problem solving, and identifying and managing side effects. Individual dietary prescriptions included the total number of calories and grams of macronutrients per day, which were divided into portions. Recipes were adapted for individual dietary prescriptions for each participant. Participants were supervised during the dietary cessation period or given further support if they chose to continue with the diet. A behavioural component to support the likelihood of adherence to the diet was incorporated, using fortnightly fidelity checklists. This was informed by the COM-B framework (17) which asserts that behaviours (in this case adherence) are influenced by three main factors; capability, opportunity and motivation. The intervention addressed these elements and drew on theories such as social cognitive theory and self determination theory.

In line with international recommendations (18) participants were advised to increase their fluid consumption and to take a multivitamin and mineral supplement that met recommended daily allowances. The modified ketogenic diet information sheet described how to manage adverse events, e.g., hypoglycaemia or hyperketosis. It was necessary for all participants to have urea and electrolytes, liver function tests, and a lipid profile prior to commencing the diet and at 6-8 week follow up to exclude significant hepatic or renal dysfunction or familial hypercholesteraemia and to identify possible adverse changes associated with the diet, such as changes in liver function or lipid profile.

### Primary and secondary outcomes

The primary outcomes of interest were participant adherence to the ketogenic diet during the study period, completion of weekly dietitian interviews, and completeness of daily ketone levels, glucose levels, and EMA data. Staff time, attrition rate, and details of side effects were also measured, and a health economics analysis was completed. A more detailed process evaluation and evaluation of the behavioural component of the intervention was carried out and will be reported separately. Detailed data on a range of secondary outcomes will also be described in a separate publication, including symptom questionnaires, physical health parameters, actigraphy, EMA data, HbA1c, C-reactive Protein (CRP), beta-hydroxybutyrate (BHB), insulin levels, fasting glucose levels, and serum and brain MRI measures of metabolites.

### Analysis

Descriptive statistics were used for recruitment, retention and outcome data completion rates. Analyses of ketone and glucose readings were performed using Excel and SPSS with appropriate measures of central tendency, frequency and dispersion. Levels of ketosis were stratified according to KetoMojo user instructions: “light ketosis” defined as 0.5-1 mmol/L, “optimal ketosis” 1-3 mmol/L and “high ketosis” 3–5 mmol/L. Three levels of hypoglycaemia were identified (19): mild (level 1; 3–3.9 mmol/L), moderate (level 2; 2.2–2.9 mmol/L) and high (level 3; <2.1 mmol/L), and percentage of occurrences calculated. Participants were considered adherent to the diet if their daily ketone levels were over 0.5mmol/L.

## Results

### Sample description

Figure 1 summarises recruitment and retention. Recruitment took place over 37 weeks, with 27 participants recruited and 26 commencing the diet. Of these 26, 8 (31%) were men and 18 (69%) were women, aged between 26-54 years (mean age 45 years). Twenty-six met criteria for type I bipolar disorder, and 1 met the criteria for type II bipolar disorder. Nine participants were within the normal BMI range, 9 were overweight and 9 were obese. Sixteen participants were prescribed an antipsychotic, 16 were prescribed a mood stabiliser and 9 were prescribed an antidepressant. Ten participants had a combination of two psychotropic classes and 4 a combination of all 3 classes. Three participants were not prescribed any psychotropic medication. Participants were from a wide geographical spread across Scotland: Aberdeen City (n=2); Aberdeenshire (n=1); Dumfries and Galloway (n=1); Dundee City (n=1); East Ayrshire (n=1); East Lothian (n=2); Edinburgh (city of) (n=8); Fife (n=1); Glasgow City (n=3); Highland (n=2); North Lanarkshire (n=2); Scottish Borders (n=1); West Lothian (n=1).

**Figure 1:**
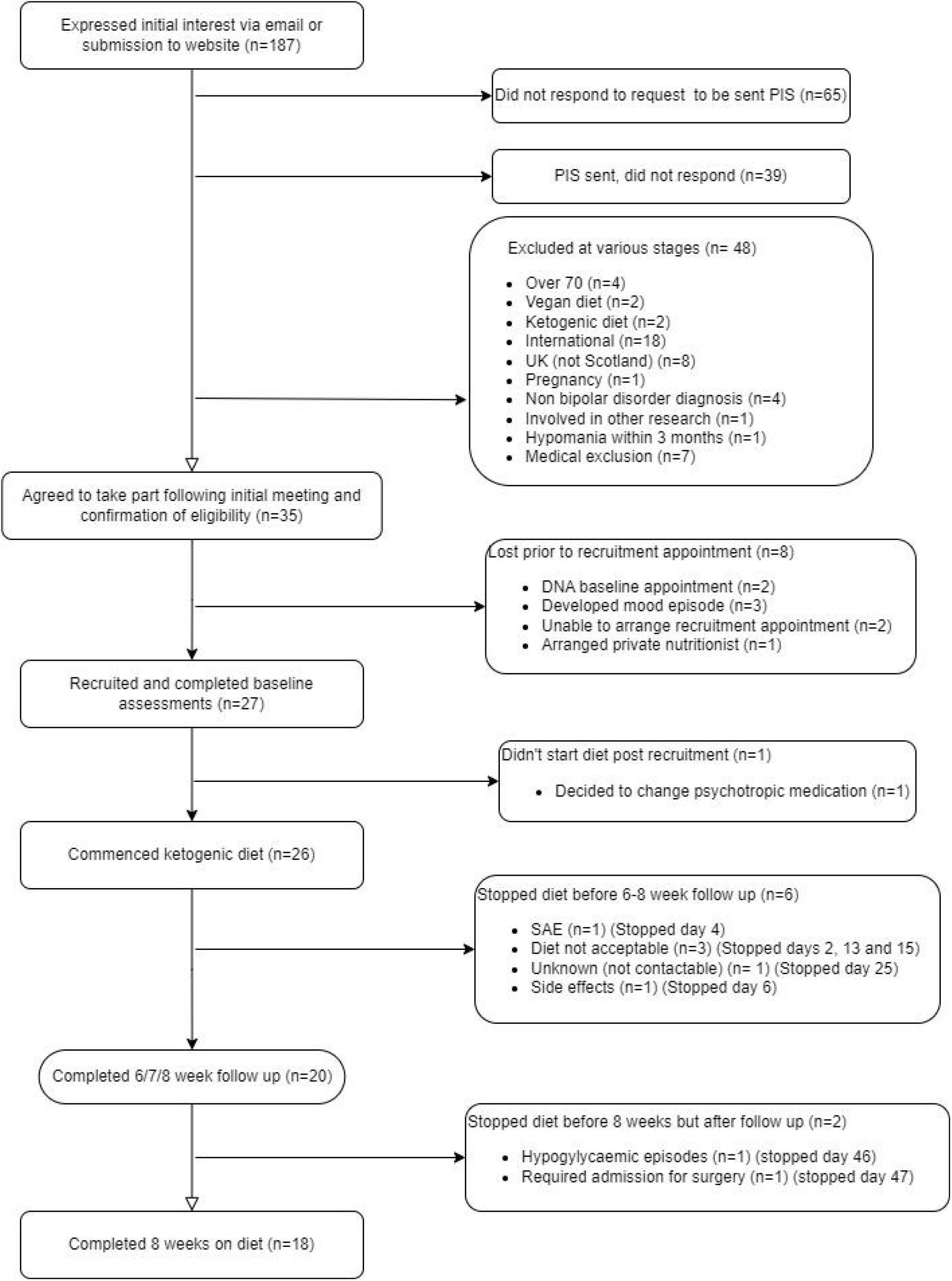
Participant flow. The flow chart illustrates the recruitment and completion rates of participants in the study. PIS, participant information sheet; SAE, serious adverse event.

Of 26 baseline appointments, 23 were in-person and 3 online. For 20 follow-up appointments 12 were in person and 8 online, excluding fasting bloods and MRI scans. Two participants completed both baseline and follow up appointments online. Twenty participants followed the diet until their 6-8 week follow up appointment, and 18 for the full 8 week intervention period. All additional contacts with the dietitian were remote and no requests for face-to-face meetings were made. All aspects of the study could be managed remotely, with no safety compromises identified.

### Data completion

Of the 20 participants who completed the intervention, 100% completed both baseline and follow-up appointments and questionnaires; 19 completed baseline and follow-up weight, height and BP; 18 completed baseline and follow-up fasting MRI scans; and 17 completed baseline and follow-up fasting blood tests. Reasons for missing assessments included one participant forgetting to fast prior to initial bloods and their MRI scan, one unsuccessful phlebotomy and one person not feeling well enough to attend follow up in person. The median time from completion of baseline appointments to starting the diet was 6 days (some participants delayed commencing the diet for practical reasons).

During the intervention period, 95% of both daily ketone and glucose measures and 93% daily EMA measures were provided by participants. In the 2-week post-intervention period this dropped to 66% for ketones, 64% for glucose and 52% for EMA data.

Forty-nine of 60 (81.7%) actigraph devices given to the 20 participants were returned with data recorded on them, with a variety of practical difficulties responsible for the missing or blank devices. The median number of days of data collected from each viable actigraph was 19 (minimum 6 days and maximum 20 days), meaning 822 of a planned 1260 (65%) days of accelerometry data were collected. One participant required a new actigraph strap as it snapped, and another reported a skin reaction to the strap, both reducing wear time. Other difficulties included postal strikes delaying participants receiving the devices.

### Physiological results

Of 45 sets of baseline and follow up blood samples from all participants, 34 demonstrated at least one parameter outside of the normal laboratory reference range that was relayed to their primary care team, the majority relating to a non-urgent altered lipid profile. All but one participant for whom baseline and follow-up data were completed lost weight during the intervention period. The median weight change was a loss of 4.5kg (9.9lbs) with weight loss ranging from 0.2 - 10kg. Weight loss was supervised by the dietitian, and intentional in some participants who were overweight or obese at recruitment, and desired weight loss.

### Daily ketone and glucose levels

The following results refer to the available ketone levels in 20 participants who continued a modified ketogenic diet for 6-8 weeks and who completed follow-up assessments. All 20 participants achieved daily readings indicating light ketosis (>0.5 mmol/L) within 1-7 days of commencing a ketogenic diet and optimal ketosis (0.5-1.0 mmol/L) within 3-13 days. Of all the readings available, 91% indicated at least light ketosis (>0.5 mmol/L). Participants’ percentage of daily readings indicating light ketosis or above ranged from 73% to 100% (Figure 2). Overall mean daily ketone levels were 1.3 mmol/L (Median = 1.1). From daily readings, the percentage indicative of ketosis was over 70% for all 20 participants (≥80% for 17 participants, ≥90% for 15 participants and 100% for 2 participants). The majority of daily glucose readings (91%) were in the normal range (4.0-7.8 mmol/L), with 9% suggesting mild hypoglycaemia (3.0-3.9 mmol/L).

**Figure 2.**
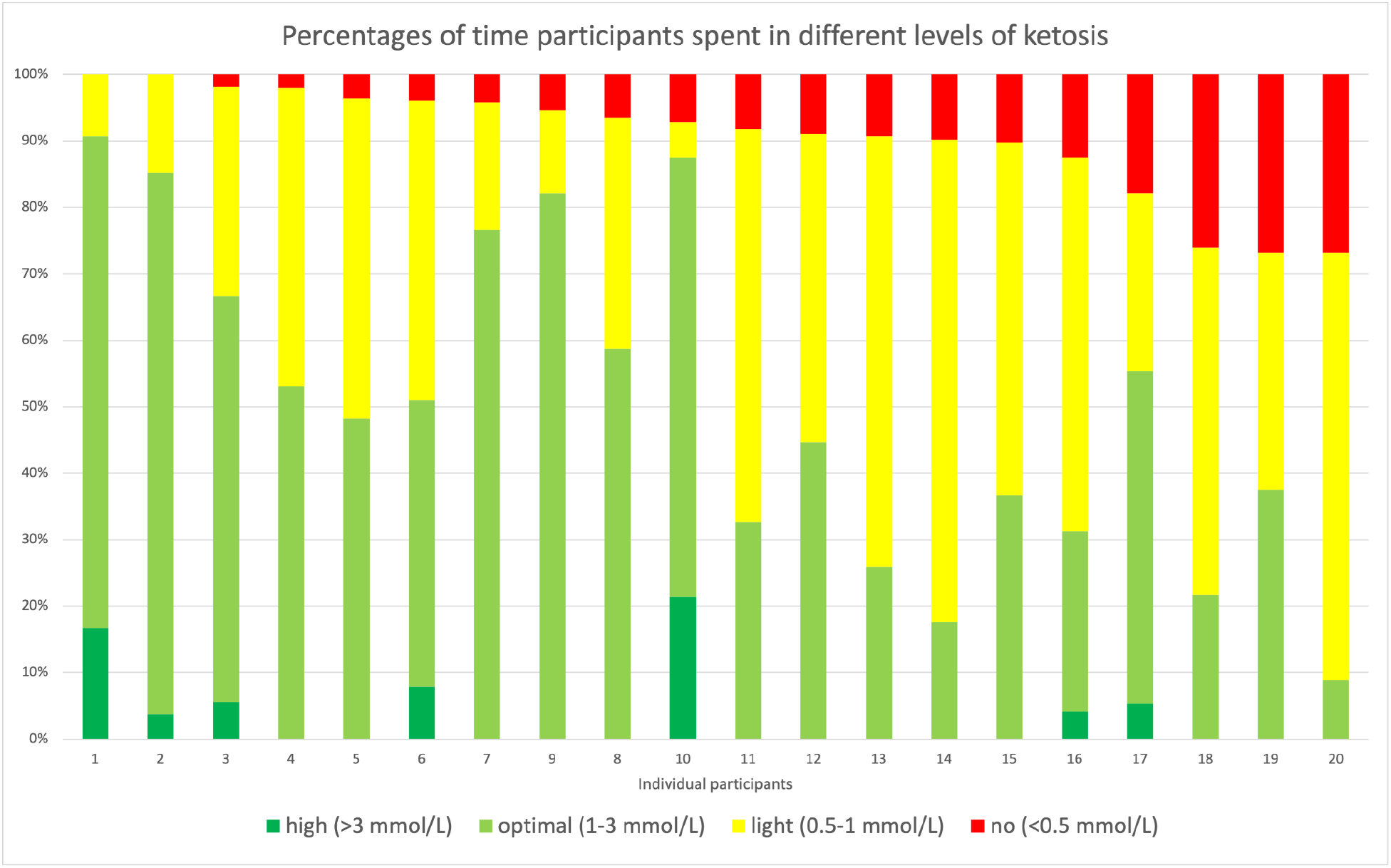
The percentage of daily participant readings representing different levels of ketosis over a 6-8 week period. Participants are ordered from highest percentage of total days with a ketone level indicating ketosis (>0.5mmol/L) to lowest. Days where data was missing were excluded from the analysis.

### Dietary changes during intervention

The study dietitian adapted and personalised the dietary prescription for each participant throughout the study. Thirty-two dietary prescription changes were required during the intervention, ranging from 0-5 per participant (Median = 1). The most frequent reasons for prescription changes were: participants preference regarding the number or size of food portions; the need to enhance ketone production; achieving prespecified loss of weight; and addressing unintended weight loss. The median dietitian time required to support each participant was 505 minutes, excluding the baseline meeting. Nine participants (45%) required a decrease in calories, ranging from 100-200 kcals, on 1-4 occasions during the intervention period as their total energy requirements had decreased following weight loss. Three participants (15%) required additional fat to promote further ketosis. Due to follow up changes in lipid profile, including an increase in total cholesterol, LDL cholesterol or triglycerides, six participants (30%) were advised to increase the amount of Omega-3 fatty acids in their diet, and to reduce saturated fat intake where possible.

### Side-effects

Table 1 shows the number and proportion of participants reporting known side-effects from a ketogenic diet, which were generally mild and resolved with dietary prescription changes. One participant had 14 incidents of mild hypoglycaemia (10 reported routinely via daily readings, 4 reported additionally), which started on day 24 of the intervention and were frequently associated with symptoms of hypoglycaemia. These continued despite 6 changes to their dietary prescription, which led to the intervention period being shortened.

**Table 2.**
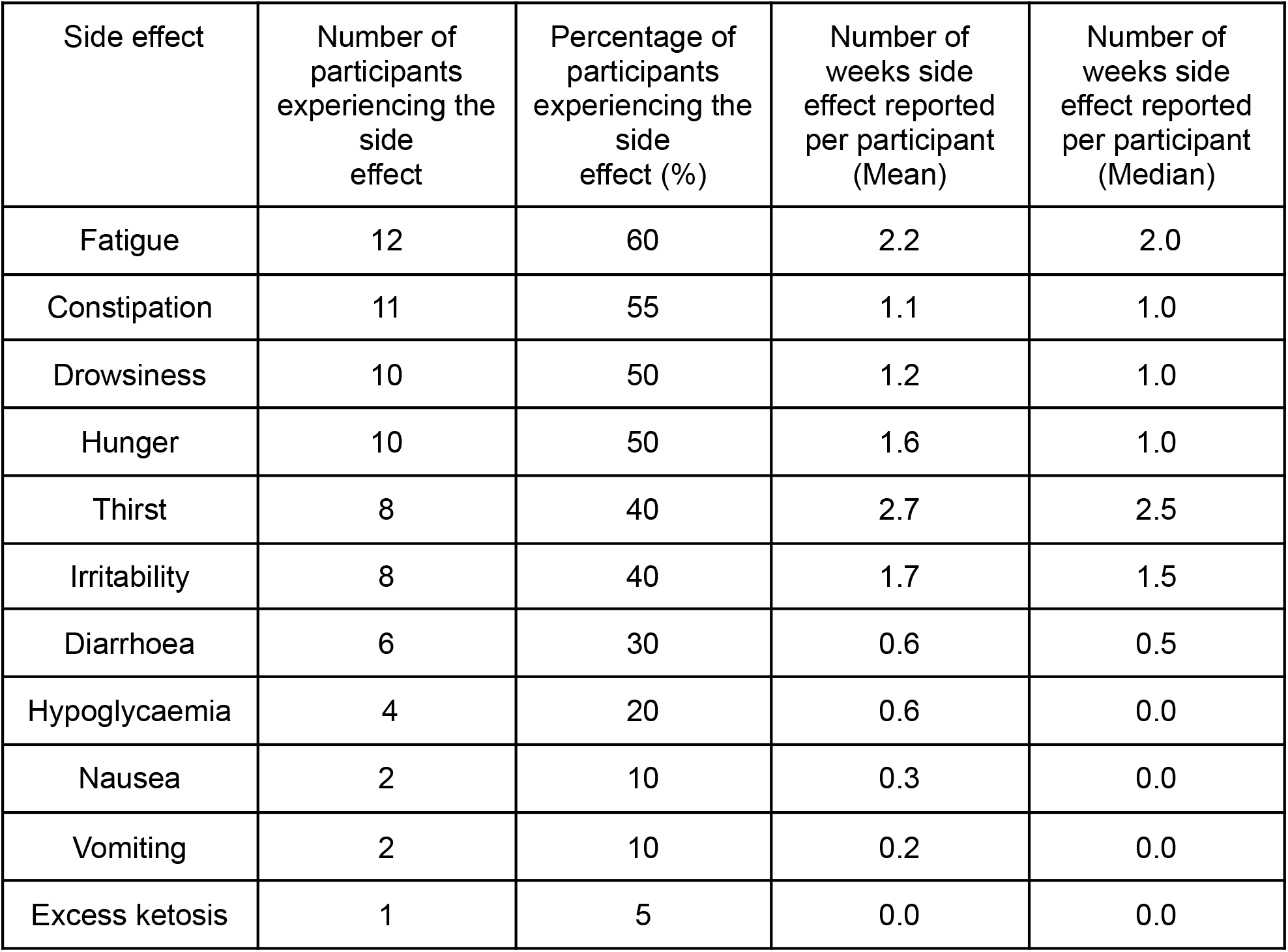
Side effects reported during the 6-8 week intervention period. The table included the data of 20 participants completing the intervention period.

| Side effect | Number of participants experiencing the side effect | Percentage of participants experiencing the side effect (%) | Number of weeks side effect reported per participant (Mean) | Number of weeks side effect reported per participant (Median) |
| --- | --- | --- | --- | --- |
| Fatigue | 12 | 60 | 2.2 | 2.0 |
| Constipation | 11 | 55 | 1.1 | 1.0 |
| Drowsiness | 10 | 50 | 1.2 | 1.0 |
| Hunger | 10 | 50 | 1.6 | 1.0 |
| Thirst | 8 | 40 | 2.7 | 2.5 |
| Irritability | 8 | 40 | 1.7 | 1.5 |
| Diarrhoea | 6 | 30 | 0.6 | 0.5 |
| Hypoglycaemia | 4 | 20 | 0.6 | 0.0 |
| Nausea | 2 | 10 | 0.3 | 0.0 |
| Vomiting | 2 | 10 | 0.2 | 0.0 |
| Excess ketosis | 1 | 5 | 0.0 | 0.0 |

### Serious Adverse Events

One participant reported ketone levels of 7.1 mmol/L on day three of the intervention and required hospital admission for the treatment of euglycemic ketoacidosis. This participant had Type II Diabetes and was prescribed empagliflozin and pioglitazone. The medical team who treated the participant in hospital concluded that this outcome was likely to be the result of combining SGLT2 inhibitors with the initiation of a ketogenic diet. Following treatment this participant recovered quickly and was subsequently withdrawn from the study.

### Barriers to following a ketogenic diet

Process evaluation data and experiences of the study team identified some potential barriers when following a ketogenic diet, including difficulty with the organisation required for food shopping and meal preparation, the affordability of food, inconvenience around the timing of meals, and increased satiety due to high fat intake. One participant had a lactose intolerance, which limited their food choices on the diet. Other process evaluation feedback included the positive impact of weight loss, improvements in energy, mood and self-control, as well as trying new foods.

### Medical input

Of the 20 participants who completed follow-up, one required a change in medication by their psychiatrist following a 1 month period of elevated mood during the study, and one reported an episode of hypomania within a week of discontinuing the diet, which lasted 3-4 weeks. One participant developed a depressive episode during the intervention period, which resolved before their follow up appointment. All study participants had some contact with a psychiatrist on the study team (NN) during follow-up, mostly to discuss administrative tasks, abnormal blood results and physical and mental health symptoms. The median contact time per participant was 18 minutes (Mean = 26).

### Health economics

Twenty participants completed all baseline and follow up health economics questionnaires, with minimal missing data (<3%). Forty-four percent of the sample were employed at baseline, and 50% at follow up. The percentage of participants reporting ‘no problems’ for each attribute in the EQ5D-5L respectively was: mobility 90% and 85%; self care 90% and 85%; usual activities 65% and 55%; pain and discomfort 45% and 45%; and anxiety and depression 45% and 50%. One participant reported level 5 (severe) for pain. The visual analogue scale (VAS) utility scores at baseline and follow up were 66.7 [Confidence interval (CI)-58.9,74.4] and 64.2 (55,73.4), respectively. Of 20 participants, almost 50% reported the effect of sickness on their work productivity and routine work. Mean productivity loss of 1.9 hours (0.83,3), in the last week was reported at baseline, and 2.4 hours (0.86,4.25) at follow-up. At baseline, mean expenditure of £14 per week (7.4,20) was reported on take away meals and £4.7 (0.1,9.2) at follow-up. Resource use was most frequently reported for GP and practice nurse visits.

## Discussion

### Recruitment, retention and mode of delivery

Overall, we found that recruiting people with bipolar disorder to a ketogenic diet intervention was feasible. Recruitment via Bipolar Scotland was successful for this pilot and could be extended to other UK-wide support groups and directly through NHS clinics for a future randomised controlled trial. Given the episodic nature of bipolar disorder, the partial loss of participants between initial and baseline meetings was expected. We also found that the majority of participants found the intervention acceptable and could be safely established and followed-up using mostly remote consultations. This approach has not been tested in other ketogenic diet studies and could facilitate recruitment from a wide geographic area in future studies.

Several additional exclusion criteria were identified during this pilot study. People with diabetes were excluded due to safety concerns after a serious adverse event in a participant taking an SLGT-2 inhibitor, and people undertaking high energy intensive exercise were excluded due to concerns about feasibility of the diet. One participant who withdrew was already engaged in intermittent fasting - future trials should consider excluding participants who regularly fast, which may increase ketone production. It is also important for researchers to have access to participant medical records during recruitment to improve the accuracy and reliability of medical information, including comorbid diagnoses.

The study withdrawal rate of 23% was comparable to other pilot studies of ketogenic diet in various conditions, ranging from 15% in recurrent glioblastoma (17), 18% in Parkinson’s disease (18) to 35% in advanced malignancies (16). The majority of withdrawals were due to difficulties with aspects of the diet (including the intensive monitoring) and not following advice to initiate gradually. To improve retention rates in a future trial, more intensive dietitian input pre-recruitment may help to increase participant understanding of what the diet and monitoring entails, including the importance of starting slowly to reduce the risk of side effects. Potential recruits could also be provided with example recipe sheets for sample ketogenic meals in advance and more detailed information on food costs. The opportunity to join group sessions before and during the study and some element of social support may also help retention (23). A more detailed qualitative assessment of participant experiences during the study will be reported separately.

We found very high completion rates of all baseline assessments, including fasting MRI scans and blood tests. One participant forgot to fast (future trials should check this before completing fasting assessments and to allow rescheduling). Despite participants needing to complete EMA data via a texting system, the overall compliance was high. Submission of ketone, glucose and EMA data was high during the intervention period but might be improved via a mobile app providing reminders. Difficulties with the completion of follow-up bloods or MRI scans were mostly due to unforeseen participant circumstances. The impact of this could be reduced by increased capacity for re-arranging blood and imaging appointments and by issuing reminders about prior fasting. The utility of the actigraphs could be improved by using more advanced technology. This could include devices with a battery life to last the entire intervention, or the ability to upload data remotely in real time. Further education regarding the benefits of collecting actigraph data may also improve compliance.

### Ketone levels and adherence

Adherence to a ketogenic diet can be challenging, and it may be expected to be especially so in bipolar disorder where depressive symptoms can have a negative impact on motivation (8). However, we found that despite the barriers identified, adherence to a ketogenic diet was high (with 91% of daily readings indicating ketosis). This was likely related to continuous support from a specialist dietitian using an individualised and adaptable approach. All of the participants were in ketosis at least 70% of the time. Weekly ketone levels from finger prick measurements showed a mean of 1.3 ± 0.8 and median of 1.1 mmol/L. These levels are similar to other ketogenic diet studies, including in Parkinson’s disease (mean daily blood ketone levels 1.15 ± 0.59 mmol/ (21)) and Alzheimer’s disease (mean weekly blood ketone levels 0.95 ± 0.34 mmol/L (24)).

There is no gold standard to measure adherence in dietary interventions but in a ketogenic diet blood ketone measurements are relatively easy and specific and more accurate than urinary levels (21). It is known that ketones levels fluctuate throughout the day, limiting the interpretation of once daily ketone measures. Continuous ketone monitoring devices may improve the accuracy of results and provide a day-to-day measure of adherence.

In future controlled trials other self-reported measures of adherence such as 24h food recalls or nutritional questionnaires could be incorporated to better understand the challenges of this dietary intervention. The production and maintenance of ketones may differ across individuals, and methods to record food intake beyond biomarker recording should increase our understanding of the barriers to achieving continuous ketosis (25).

### Side effects and adverse events

Side effects were common and typically mild and could usually be resolved with dietary adjustments such as increased fibre content and fluid intake for constipation (18). These were uncontrolled observations and so it is also not possible to determine whether each adverse event was directly related to the intervention. Although there were changes in lipid profile (increased levels of total cholesterol, LDL or triglycerides) at follow-up in 9 participants, this was expected and usually returns to normal limits over time (26, 27). Four participants had one or more incidents of low blood glucose levels (<4 mmol/L) but these resolved with dietary changes, highlighting a need for regular monitoring. One participant had persistent mild but symptomatic hypoglycemia and struggled to maintain a blood glucose of >4 mmol/L despite many dietary changes, indicating the need to consider stopping the diet if this occurs.

The serious adverse event in the form of euglycemic ketoacidosis during dietary initiation was a novel finding. Other trials using a ketogenic diet as an intervention have started excluding participants on SGLT2 inhibitors, which is important for future studies.

### Health Economics

Health economic instruments were completed with minimal missing data, indicating their feasibility for future trials in this population. The EQ5D-5L VAS mean score of 64-68 can be benchmarked against population values of 83-85 (depending upon the source used), indicating a lower quality of life in this participant group. The findings from the health economic data are indicative of the level and variability of all reported values, possible changes in baseline levels of expenditures, quality of life, and productivity at follow up; and applicability of using such health economic instruments larger scale studies.

### Strengths and limitations

The strengths of our study include relatively high completion of baseline and follow-up assessments including detailed resource use and quality of life instruments required for any future cost-effectiveness analyses, high completeness of the ketone, glucose and EMA reading, successful induction and maintenance of ketosis and effective delivery of dietetic support. The identification of new exclusion criteria will improve the safety and efficacy of a future trial. Despite the barriers identified, 77% of participants remained on the diet until follow-up, consistent with other ketogenic diet research (28).

The nature of the ketogenic diet used required a cohort of participants with a high level of motivation, commitment and time. This may limit the applicability of this intervention to the wider bipolar disorder population. We also limited our study to people who were currently euthymic. In the future it would be helpful to assess the feasibility of this intervention for bipolar depression.

## Conclusions

This pilot study is one of the first to explore the feasibility and safety of a ketogenic diet in people with bipolar disorder. We provide evidence that this intervention is feasible and safe. Moreover, it is clear that people with bipolar disorder are open to considering dietary/nutritional interventions as adjunctive therapies and that the predominantly remote recruitment and conduct of a ketogenic diet intervention is possible. Our study also showed that a future trial could readily identify and measure all the resource use, quality of life and wider household and productivity impacts relevant for conducting a full cost-effectiveness/cost-utility analysis. We conclude that the next step should be a randomised controlled trial of a ketogenic diet in bipolar disorder.

## Data Availability

The data that support the findings of this study are not publicly available as explicit consent was not sought from participants.

## Author Contributions

IC, HC, and DS were responsible for conceptualising the research question. NN, IC, HG, SS, EM, TM, KB, MT, MM-G, JN, AM, CF, HC and DS were responsible for designing the study. NN, HG, BM, FC and BR were responsible for data collection. NN, IC, HG, EM, PB and IK were responsible for analysis and interpretation of the data. NN, HG and IK were responsible for drafting the paper. All authors were involved in subsequent revisions, and agree to be accountable for all aspects of the work.

